# Displacement ventilation: a viable ventilation strategy for makeshift hospitals and public buildings to contain Covid-19 and other airborne diseases

**DOI:** 10.1101/2020.04.22.20075648

**Authors:** Rajesh K. Bhagat, P. F. Linden

## Abstract

The SARS-CoV-2 virus has so far infected more than 2.4 million people around the world, and its impact is being felt by all. Patients with airborne diseases such as Covid-19 should ideally be treated in negative pressure isolation rooms. However, due to the overwhelming demand for hospital beds, patients are being treated in general wards, hospital corridors, and makeshift hospitals. Adequate building ventilation in hospitals and public spaces is a crucial factor to contain the disease^1,2^, to exit the current lockdown situation, and reduce the chance of subsequent waves of outbreaks. Lu et al. ^3^ reported an air-conditioner induced Covid-19 outbreak, by an asymptomatic patient, in a restaurant in Guangzhou, China, which exposes our vulnerability to future outbreaks linked to ventilation in public spaces. We demonstrate that *displacement ventilation* (either mechanical or natural ventilation), where air intakes are at low level and extracts are at high level, is a viable alternative to negative pressure isolation rooms, which are often not available on site in hospital wards and makeshift hospitals. Displacement ventilation produces negative pressure at the occupant level, which draws fresh air from outdoor, and positive pressure near the ceiling, which expels the hot and contaminated air out. We acknowledge that, in both developed and developing countries, many modern large structures lack the openings required for natural ventilation. This lack of openings can be supplemented by installing extract fans. We provide guidelines for such mechanically assisted-naturally ventilated makeshift hospitals, and public spaces such as supermarkets and essential public buildings.

## Introduction

Covid-19, the disease caused by the virus, SARS-CoV-2 is a pandemic and a global emergency which has stressed and in some cases overwhelmed healthcare systems across the world^4^. As of April 20, 2020, more than 2.4 million confirmed cases and over 165,000 fatalities have been reported, and these numbers are expected to increase significantly. Originating in Wuhan in China, it has rapidly spread to most of the world, and the current epicentre of the disease is in Europe and the United States. Outbreaks of the disease have already occurred in high population density, low income/developing countries such as India, Pakistan, Nigeria, and at some later stage, the disease is expected to cause significant numbers of fatalities in these countries as well.

The proliferation of the disease has overwhelmed many existing healthcare systems. In response to the overwhelming demand for hospital beds for critically ill patients, a new healthcare facility/hospital has been built from scratch in record time in Wuhan. Similar healthcare preparedness is imperative in both the developed and the resource-scarce developing world. In London, an exhibition and convention centre has been converted into a 4000-bed Nightingale-style emergency hospital in nine days. Other such new emergency hospitals are being created in the UK in Manchester, Birmingham, Bristol, and Glasgow. In New York, makeshift tent-hospitals have been created in the Central Park, and the Manhattan convention centre has been converted into a makeshift hospital. Suspected Covid-19 patients are being quarantined in hotels, and in India railway coaches are being converted to accommodate the patients and suspected cases.

Covid 19 is an infectious respiratory disease. The virus transmits through direct or indirect human to human contact; fomite and respiratory droplets, respectively^5^. The virus can remain stable in aerosol form for hours^6^, and can still potentially infect people^4,7,8^. In 2003, a possible SARS-CoV infection to healthcare workers during an aerosol-generating medical procedure had been reported^9^. Furthermore, Yu et al. ^10^ presented the evidence of long-distance airborne SARS contagion transport between the apartment blocks in Amoy Gardens housing complex in Hong Kong.

Conventionally, patients with infectious respiratory diseases are kept in negative pressure isolation rooms, which provide ventilation to control the spread of the disease. However, in the current scenario patients are kept in the hospital wards and beds which were not meant to handle such cases. In this paper we show that *displacement ventilation*, either natural or mechanical where air is drawn in through low-level inlets and extracted at high level, can significantly reduce the spread of airborne contagion, and save the lives of healthcare workers, caregivers, and patients^5^. After the end of the current lockdown, this easily implementable ventilation strategy, can be used in other public building, schools, restaurants etc., to reduce the chances of any subsequent outbreaks, and improve public life.

### Building ventilation in healthcare settings

The purpose of ventilation is to remove harmful and unwanted agents – gases, pollutants, heat, and contagion from the building envelope by indoor-outdoor exchange of air. There are two basic ventilation types (Figure 1); *mixing ventilation* where the air is circulated throughout the whole space, for example by ceiling fans or cool air introduced from high-level airconditioning ducts with a view to maintaining uniform conditions such a temperature throughout the space, and *displacement ventilation*, where fresh air is introduced through low-level inlets and extracted at high level in the space. Of these two modes mixing ventilation is the most commonly used, both in mechanical and natural ventilation.

**Figure 1:**
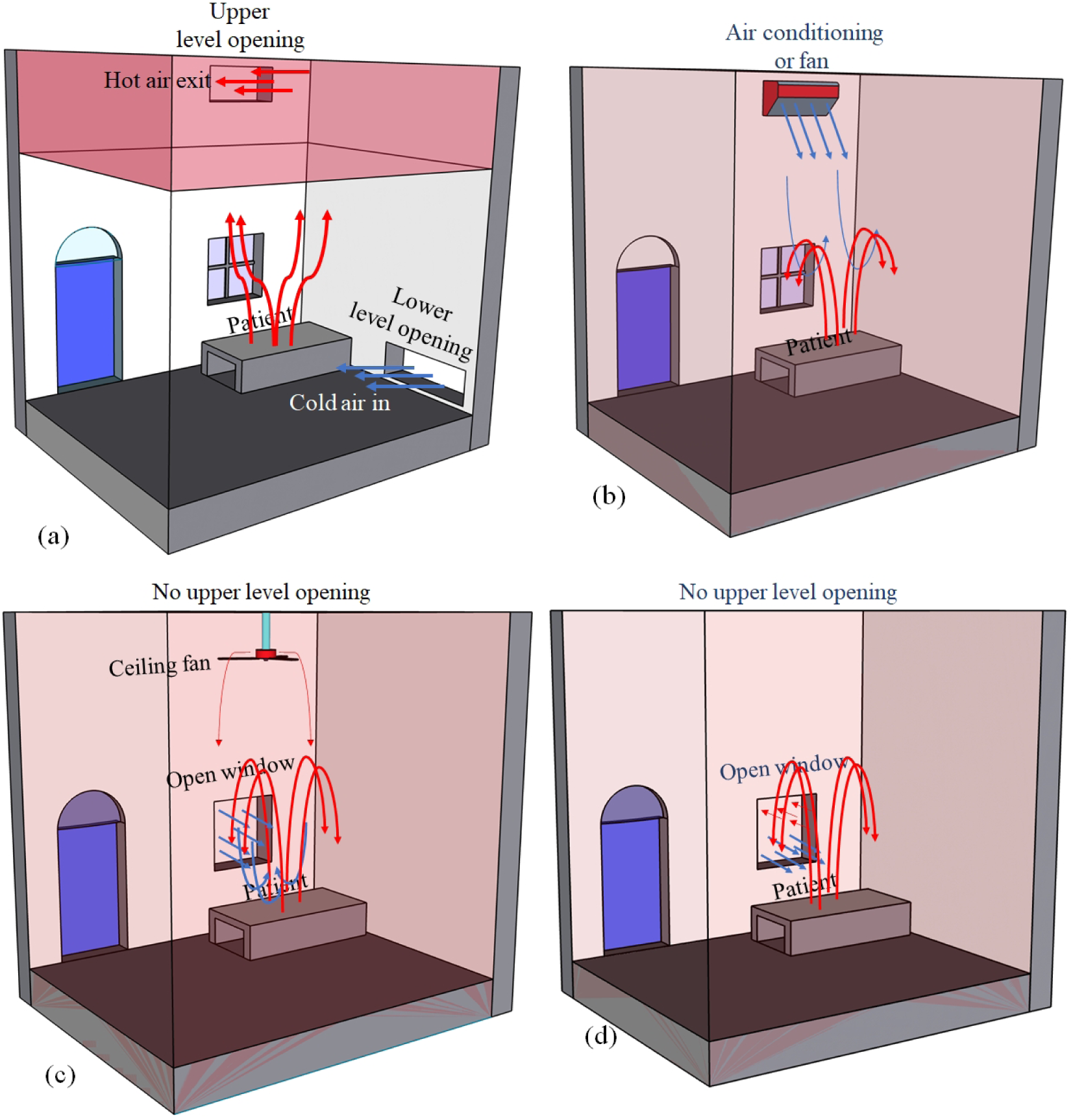
Schematic of different ventilation types, (a) displacement ventilation, (b) positive pressure mixing ventilation by air-conditioning, (c) positive pressure mixing ventilation by fan and open window, and (d) wind-driven mixing ventilation. In displacement ventilation fresh air enters from the bottom of the room and contaminated air exits from the top. In mixing ventilation with a ceiling fan or high level air conditioning unit, the contaminant/contagion is distributed uniformly throughout the whole room. These ventilation modes can either be produced mechanically by fans or air conditioning, or by using natural ventilation.

Displacement ventilation relies on the fact that inevitably heat is generated within the space, either from the occupants and/or equipment, and this hot air rises and accumulates near the ceiling. Consequently, the ambient conditions experienced by occupants in these two different ventilation modes are fundamentally different. In mixing ventilation the occupant is surrounded by air of uniform properties, such as the concentration of a contaminant – a bit like lying in dirty bath water. In contrast, in displacement ventilation the occupant is surrounded by newly arrived air and pollutants generated near or by the person are lifted towards the ceiling by the rising warm air – rather like being in an inverted shower and rinsed with clean water. Of course, for the same amount of contaminant generation and ventilation flow the total amount of contaminant in the space is the same: in mixing ventilation it is everywhere at a relatively low concentration, while in displacement ventilation it is concentrated in a warm layer near the ceiling. When the system is designed correctly this contaminated layer is located above the occupants and so they are only exposed to clean uncontaminated air.

Consequently, displacement ventilation will provide much better local conditions for patients within hospitals. How is this achieved? Essentially fresh air needs to be introduced at low level in the space so that it flows over the patient collecting air expelled from her lungs and heat released from her body and any surrounding equipment. This air is then warmer and rises above the patient and accumulates near the ceiling where it needs to be extracted from the building. As a consequence of this efficient flow pattern, the ventilation rate, i.e. the rate at which air is exchanged between inside and outside a space, required for a healthy environment in displacement ventilation is much less than that required for mixing ventilation (see figure 2). For contagious disease hospital wards, the World Health Organisation, WHO recommends – apparently based on mixing ventilation – a ventilation rate of 160 LSP (Litres per Second per Person). This is much greater than the 10 LSP of fresh air required for breathing^11^, and would require 60 air changes per hour (ACH) in a typical cubicle measuring 4 m wide, 3 m long and 3 m high. This value is much higher than experienced in most hospitals which have ACH of 12-15 at most^12^. For patients with an airborne contagious disease, a negative pressure ventilated isolation room is ideal, as it does not allow contagion to escape from the isolation room. However, building and running new isolation rooms requires time, resources, and maintenance staff, which in the current scenario are scarce. To contain the disease, displacement ventilation in makeshift hospitals and other buildings can be a viable and safe alternative.

**Figure 2:**
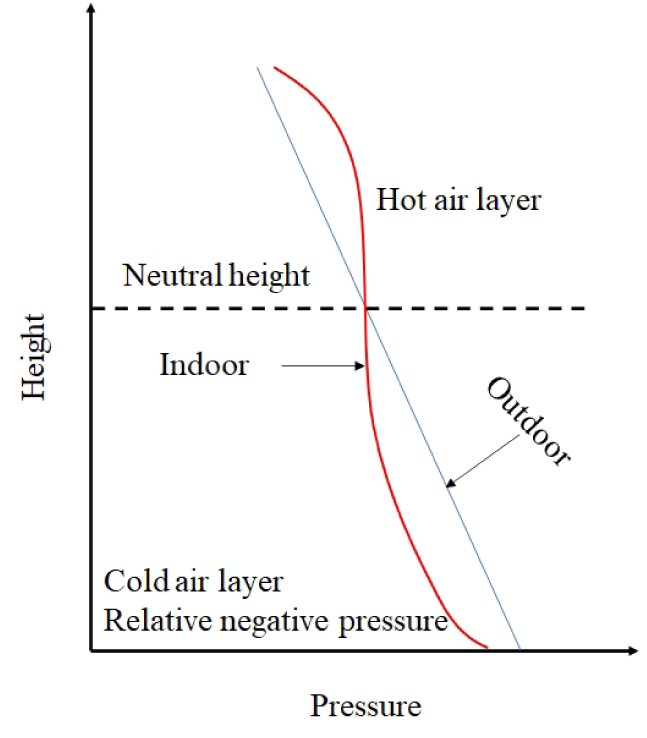
Pressure distribution, and neutral height inside a naturally ventilated building. Below the neutral height, indoor pressure is less than the outdoor pressure at the same level, producing a negative pressure at low level that sucks in outside air. The positive inside pressure at high level drives warm polluted air out through the upper openings. (Figure adopted from Linden ^15^).

Mixing ventilation, as the term implies, mixes the air within a space. This implies that aerosols and other contaminants are mixed throughout the space so that all occupants are exposed to them. In the case of Covid-19 the virus is transported on droplets expelled from an infected person. The exact nature of this transport depends on the distribution of droplets, whether the individual is coughing, etc. and is beyond the scope of this paper. However, Lu et al. ^3^ reported an air-conditioning promoted Covid-19 outbreak in a restaurant in China, where one asymptomatic Covid-19 positive person infected other occupants in the room who were more than 2 m away. Indeed, in enclosed spaces such as hospital wards mixing caused by air conditioning or ceiling fans spreads the contamination throughout the space and so makes the ‘2 m’ social distancing rule irrelevant. Instead we recommend that displacement ventilation be used to provide a safe environment for occupants in a building. We next discuss displacement ventilation in two contexts, natural ventilation and mechanical ventilation.

### Natural ventilation

Both modes of ventilation shown in figure 1 can be produced either mechanically or by natural ventilation. Natural ventilation relies on free natural resources, temperature differences between inside and outside the building – known as ‘stack-driven’ ventilation and the wind, to drive flow through the building. There is evidence that natural ventilation can be beneficial for patient health even when mechanical ventilation is available^1,2^, and in many cases it will be the only option.

#### Wind-driven natural ventilation

Wind-driven ventilation depends upon, wind speed and direction and turbulence intensity, all of which fluctuate in time. Urban layout, building type, building location, the size and density distribution of the surrounding buildings, and the urban heat island, make it extremely challenging to predict the ventilation rate. Furthermore, the fluctuating indoor flows caused by wind driven ventilation may produce random distribution of the unwanted agents, which in the context of removal of the airborne contagion from patients is undesirable. However, when openings are appropriately arranged, for example by ensuring that roof extracts are at negative wind pressure, wind-driven ventilation can assist displacement ventilation and make it more effective^13^. Consequently, in the following we ignore the effects of wind and concentrate on stack-driven ventilation, noting that care must be taken to ensure that any wind-driven flow assists and does not disrupt the flow patterns established by temperature differences.

#### Displacement ventilation or stack-driven natural ventilation

In stack-driven natural ventilation warm, buoyant air (due to body heat or the heat generated due to appliances) rises towards the ceiling and exits through an upper level opening. This in turn draws in cooler outdoor (high density) that flows across the floor of the room. The stratification produced by the indoor temperature gradient drives the flow inside the building (figure 2). The flow is predominantly unidirectional, upwards from the floor, removing airborne contagion away from patients and other occupants towards the ceiling where it gets flushed out of the building. Particles and aerosols generated indoors by the occupants due to breathing and other activities rise with the warm air and are trapped in the hot upper air layer, which is advantageous in hospitals^14^.

### Design guidance for a hospital: natural ventilation

We first calculate how much openable area needed to ventilate a space. The effective openable area *A*\* in the façade of a building needed to provide a unpolluted lower layer of height *h* in a room of height *H* is given by

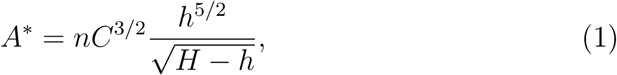

where *n* is the total number of occupants (we can assume the occupants to be equal strength heat sources), and the empirical constant *C*≈ 0.105 (Linden ^15^ & Morton et al. ^16^). This effective openable area *A*\* depends on a combination of the total areas *a*_*t*_ and *a*_*b*_ of the top and bottom openings, respectively, given by the relation

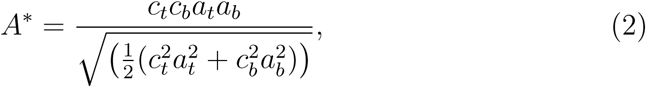

where *c*_*t*_ ≈0.65 is the discharge coefficient of the top opening, and *c*_*b*_ ≈ 0.6 is the pressure loss coefficient for the bottom opening. For given values of the height of the space *H* and the number *n* of occupants, (1) and (2) can be solved to find the areas *a*_*t*_ and *a*_*b*_ required to ensure the occupants remain in the unpolluted layer of height *h*.

As an example, suppose that a patient and two nurses (i.e. *n* = 3) are in a bay with a total height *H* = 4 m. To ensure the unpolluted layer is higher than 2 m, (1) implies that *A*\* ≈ 0.4 m^2^. If the top and bottom areas are equal in size, this requires that they are both approximately 0.64 m^2^, i.e. they measure, say, 800 mm by 800 mm. It may be difficult to achieve this amount of opening in practice and so some mechanical extraction may be necessary. We address this possibility in the next section.

When designed properly, the polluted upper layer should be above occupants heads. From (1), it is clear that buildings with tall ceilings and with large upper level and lower level openings are optimal for naturally ventilated makeshift hospitals.

### Design guidance for a hospital: mechanically-assisted natural ventilation

Buildings are generally equipped with large lower level openings (windows and doors) but often lack large upper level openings. As mentioned above, shows that taller spaces are better for natural displacement ventilation – *A*\* decreases as *H* increases implying that smaller openings will suffice in a taller space. However, in situations where the required opening area is not available or the space is not tall enough, natural ventilation can be supplemented or replaced by mechanical extraction from the upper part of the space.

In this case the height *h* of the lower clean zone is determined by matching the mechanical extraction rate *Q* with the flow of warm air from the occupants etc. into the upper warm zone. This is given by Linden et al. ^17^ by the formula

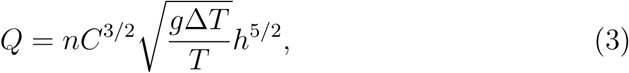

where *g* = 9.81 m s^*−*2^ the acceleration due to gravity, *T* is the outside temperature (in Kelvin) and *T* +Δ*T* is the temperature of the warm upper layer.

Note now that the height of the space is no longer important and the depth of the clean zone is set by the extraction rate.

Consider again the case of a patient and two nurses and suppose the outside temperature *T* = 293 K, (20 ^*°*^C) and the inside warm layer is at *T* + Δ*T* = 303 K, (30 ^*°*^C). Then to attain *h* = 2 m, *Q* = 0.33 m^3^ s^*−*1^. This corresponds to approximately 110 litres per second per person, 30% less than the WHO guideline for mixing ventilation. Note that in hot climates, cool air can be introduced at low level using underfloor air distribution rather than through overhead air conditioning ducts ^18^.

### Guidelines for displacement ventilation of makeshift hospitals

- Tall rooms with large upper and lower level openings are suitable for makeshift hospitals.
- Any upper-level ceiling fans or air conditioning units should be removed or not used. Underfloor air distribution should be used if air conditioning is employed ^18^.
- The lower unpolluted layer height should at least be 2 m deep, and for a building equipped with upper and lower level openings (naturally ventilated), the maximum number of occupants can be calculated by solving (1) and (2).
- To supplement the unavailability of openings, extractor fans (mounted near the ceiling) could be used. For a given space, the number of occupants, and unpolluted layer height, the extraction flow rate can be calculated from (3).
- Except in open-plan hospitals, individual cubicles should be equipped with local lower-level openings. Warm air extract can be centrally arranged.
- In hot climates with uncomfortable outdoor air temperature, cold air should be supplied near the bottom of the room, and hot air exit vents should be made near the ceiling.

## Conclusions

Adequate ventilation is essential to maintain a healthy indoor environment in hospitals and other public buildings. In addition to the ventilation rate, the dominant flow pattern is an important factor for the removal of contagions, and cross-contamination between the occupants (patients, healthcare staffs, and caregivers). The commonly used mixing ventilation tends to enhance the spread of airborne pathogens^3^. With the current practice at hospitals, where both the staff and patients both wear adequate PPE (masks, respirators, etc.), displacement ventilation (natural or mechanical) is a suitable and easily implementable alternative to the negative pressure ventilation for makeshift hospitals. Displacement ventilation is economically and environmentally sustainable, and after minor retrofitting, it can easily be implemented into existing buildings.

In order to exit from the current situation and to prevent secondary outbreaks, building ventilation strategy must play an important role. After the end of the lockdown, we believe that public life and practices will change, and persons displaying symptoms will be discouraged from attending public gatherings. Displacement ventilation in public spaces will provide additional protection from asymptomatic patients, and improve public life. Therefore, we recommend that public spaces (bars, restaurants, supermarkets, etc.), should be retrofitted for displacement ventilation to minimise further spread of the disease after lockdown.

## Data Availability

The manuscript contains no data.

## Acknowledgements

We are grateful for helpful comments on this article from Sophy Bristow, Megan Davies Wykes, Shiwei Fan and Anna Schroeder. This work was supported by the UK Engineering and Physical Sciences Research Council (EPSRC) Grand Challenge grant ‘Managing Air for Green Inner Cities (MAGIC) grant number EP/N010221/1.

## Author’s contributions

Both authors contributed equally to the content and writing of the paper.

